# Biofluid Biomarkers of Ischaemic Penumbra in Acute Ischaemic Stroke: A Systematic Review and Meta-Analysis

**DOI:** 10.64898/2026.03.03.26347352

**Authors:** Yuki Kawamura, David S Liebeskind, Shubham Misra, Erum I Khan, Amr Elshahat, Pei Yi Chook, Ethan Wang, Mark Reed, Melissa C Funaro, Priyal Tiwari, Adam de Havenon, Charles R Wira, Tu Kiet Lam, Srikant Rangaraju, Maarten G Lansberg, Nishant K Mishra

**Affiliations:** Department of Neurology, Yale School of Medicine, New Haven, CT, USA; School of Clinical Medicine, University of Cambridge, Cambridge, UK; Department of Neurology, University of Southern California, CA, USA; Department of Radiology and Biomedical Imaging, Yale School of Medicine; Universiti Malaya, Kuala Lumpur, Malaysia; College of Medicine, California Northstate University, CA, USA; Harvey Cushing/John Hay Whitney Medical Library, Yale University, New Haven, CT, USA; Palliative Medicine, Sir Ganga Ram Hospital, New Delhi, India; Department of Emergency Medicine, Yale University School of Medicine, New Haven, CT; Keck MS & Proteomics Resource, Yale School of Medicine, New Haven, CT, USA; Department of Neurology – Division of Vascular Neurology, Stanford Hospital, Palo Alto, CA

**Keywords:** Ischaemic Stroke, Penumbra, Biomarkers, Blood, Biofluid, Humans, Animals

## Abstract

**Background and Objectives:** The ischaemic penumbra is the principal therapeutic target in acute ischaemic stroke (AIS). Although perfusion imaging identifies salvageable tissue, its availability is limited and iodinated contrast exposure carries risk. Validated blood-based biomarkers could serve as scalable surrogates for imaging-defined penumbrae. We conducted a systematic review and meta-analysis assessing associations between blood-based biomarkers and ischaemic penumbrae.

**Methods:** We searched MEDLINE, Embase, PsycInfo, Web of Science, and Cochrane Central Register of Controlled Trials until December 3, 2025 for studies involving human (≥18 years) or animal subjects with AIS reporting the presence of ischaemic penumbra. The primary outcome was the difference in mean biomarker levels in subjects with and without penumbrae as defined by the study authors. Risk of bias was assessed using QUADAS-2. We calculated each biomarker’s pooled standardized mean difference (SMD) and 95% CI where possible. Protein-protein interaction (PPI) network and pathway analyses were conducted using Cytoscape and enrichR (PROSPERO: CRD42023453175).

**Results:** We identified 11 studies (1546 human subjects and 11 animals) that assessed 53 candidate biomarkers in humans and 2 in animals. Two studies had a low risk of bias, while nine had concerns. Meta-analysis of six biomarkers (three studies) identified two biomarkers demonstrating significant association with presence of a penumbra in humans: interleukin-10 (IL-10; SMD 2.40 [1.37 to 3.42]) and interleukin-6 (IL-6; SMD -0.93 [-1.80 to -0.06]). However, substantial heterogeneity (I² >90%) limited confidence in effect size precision. Of the 47 human biomarkers that had insufficient data for meta-analysis, 38 showed significant association with ischaemic penumbrae. ORAC_PCA_ (SMD 0.31 [0.01 to 0.60]) and MR-proADM (SMD 0.97; [0.52 to 1.42]) were higher in the penumbra group, and 35 RNA biomarkers (e.g., IL1B r = -0.59, p = 0.003; circOGDH r = 0.962, p = 0.002), and NT-proBNP (r = 0.199, p < 0.001) significantly correlated with penumbra volume. Pathway enrichment revealed positive associations with angiogenesis and IL-12 signalling, and negative associations with leukocyte migration, chemokine signalling, and platelet activation.

**Discussion:** Multiple genes and proteins show promise as biomarkers of the ischemic penumbra, but most have not been prospectively validated. Pathways implicated by these biomarkers converge on inflammatory regulation, haemostasis, and cerebral perfusion. Rigorous prospective validation is required before integration into prehospital or emergency triage workflows.

## Introduction

A key consideration in decision-making for initiating endovascular therapy in AIS is the presence of an ischaemic penumbra, the area surrounding the core in which blood flow is reduced but neuronal recovery is possible if blood flow resumes^1^. Radiologically, the penumbra is identified by the mismatch between the ischaemic core and perfusion deficit, indicating brain tissue at risk. Salvaging the penumbra tissue can minimise neurological deficits after a stroke.

Thrombolytic therapy in AIS is underutilised due to a 4.5-hour window^2, 3^, although accumulating evidence supports use beyond this window with advanced neuroimaging^4–9^. Eligible patients may receive endovascular therapy up to 24 hours after symptom onset particularly when a significant mismatch is present, as patients with a mismatch tend to have better outcomes after reperfusion therapy^10, 11^. Thus, perfusion imaging aids selection of patients with salvageable penumbrae, but it may not always be available. In a US survey (2020-2021), CT perfusion imaging (CTP) was available in 39% of emergency departments overall and 23% in those at hospitals with low stroke volume^12^. Similarly, 30.1% of stroke patients received CTP in a Korean study (2018-2021)^13^, indicating that the availability of advanced neuroimaging can be a constraint even in resource-rich countries.

Biofluid biomarkers could offer a cost-effective method for triaging patients at the point of care and have clinical utility in reducing thrombectomy delays, which is associated with worse outcomes^14^, and in avoiding unnecessary contrast use. Yet, current evidence on these biomarkers is limited and not systematically studied. We conducted this systematic review to evaluate studies on biofluid markers of ischaemic penumbra and to analyse their association with pathogenic mechanisms.

## Methods

### Information Sources

MEDLINE, Embase, PsycInfo, Web of Science, and Cochrane Central Register of Controlled Trials databases were searched on September 6, 2023 and December 3, 2025. References were uploaded to Covidence for screening and duplicate references were removed. Search strategies were developed in consultation with a librarian (M.C.F.) and are reported in the supplementary material.

### Eligibility Criteria

We included studies with human subjects aged over 18 years and animal subjects with AIS and ischaemic penumbra confirmed by mismatch on neuroimaging that reported any fluid biomarker measurements. Given substantial variability in penumbra definitions across studies (radiographic mismatch thresholds, clinical–diffusion mismatch, and perfusion-core ratios), we accepted study-specific definitions but accounted for this heterogeneity in interpretation of pooled analyses. We included studies treating the presence of penumbrae as a dichotomous variable as well as those which correlated biomarker concentrations with the size of penumbrae. Both radiological and clinical-radiological mismatches were included, in accordance with current criteria for mechanical thrombectomy^15^. No restrictions were applied based on date, language, gender, or ethnicity. We excluded studies that focused solely on haemorrhagic stroke and lacked full texts, reviews, conference proceedings, preprints, case reports, case series, or ongoing/unpublished studies. The protocol was prospectively registered on PROSPERO (CRD42023453175).

### Outcomes

Our primary outcome was the difference in mean biomarker levels in subjects with and without ischaemic penumbra. Our secondary outcome was the correlation between biomarker levels and penumbral volume.

### Data extraction

The systematic review was conducted in accordance with the 2020 PRISMA reporting guidelines^16^ (Supplementary material). Nine reviewers (Y.K., S.M., P.Y.C., E.W., M.R., N.P, A.E., A.M., and E.K.) independently screened the titles and abstracts for eligibility, as well as for duplicate studies, after which full texts were screened for inclusion. Conflicts were resolved by discussion with a senior author (N.K.M.). Subsequently, the following information was systematically extracted from each eligible study: study author surname, publication year, sample size, model organism, age, sex ratio, hypertension, smoking, atrial fibrillation (AF), prior stroke, diabetes, dyslipidaemia, study design, mismatch calculation method, mismatch volume, definition of mismatch presence, type of neuroimaging used, ischaemic core volume, aetiology (TOAST classification), NIH stroke scale (NIHSS), presence of reperfusion therapy, time from stroke to fluid collection, biomarkers assessed, biomarker type, biomarker detection method, biofluid type, biomarker levels, threshold value for prediction, AUC, sensitivity, specificity, and odds ratio. Study authors were emailed twice to obtain additional information when insufficient data were available to conduct the analysis.

### Risk of Bias (Quality) Assessment

The quality of the studies was assessed by two independent reviewers (E.K. and A.E.) using the Quality Assessment of Diagnostic Accuracy Studies-2 (QUADAS-2) framework^17^, a 13-point tool comprising four domains for bias assessment and three for applicability. Discrepancies were resolved by discussion with the senior author (N.K.M.). Studies scoring “low” in all domains were deemed to have a low risk of bias or low concern regarding applicability, whereas studies with one or more domains scoring “unclear” or “high” were deemed to be at risk of bias or having concerns regarding applicability, according to the QUADAS-2 recommendations^17^.

### Statistical analysis

Biomarker levels were calculated as mean and standard deviation for the cases (subjects with penumbra) and controls (subjects without penumbra), where dichotomous results were available directly from the studies or using conversion formulae^18^. For each biomarker, standardised mean differences and 95% confidence intervals (CI) were calculated after pooling studies where applicable. A meta-analysis was performed for biomarkers featured in more than one study using the “meta”, “metafor”, and “dmetar” packages in R version 4.3.2. Heterogeneity was calculated using the I^2^ statistic, and characterised as low if I^2^ <25%, moderate if I^2^=25-75%, and high if I^2^>75%^19^. Different cohorts within the same study were combined unless they were non-overlapping and assessed different definitions of penumbra. A random-effects model was applied for pooled analyses given anticipated clinical and methodological heterogeneity across studies; fixed-effects estimates were explored in sensitivity analyses where heterogeneity was low. Correlation coefficients are presented exactly as reported in the studies.

### Protein-protein interaction network analysis

Protein-protein interaction (PPI) network analysis was performed using the Cytoscape 3.10.3^20^ software using the STRING 12 database^21^ to query for known protein-protein interactions. A network was generated with biomarkers as nodes and interactions as edges, with the number of interactions illustrated as the colour of the nodes and the strength of interaction as the thickness of the edges.

### Pathway analysis

Protein biomarkers were converted to their RNA transcript names, and over-representation analysis was performed with the enrichR package^22^ using the Reactome, KEGG, and GO databases. Pathways were included only if more than 3 genes mapped to the pathway and if biomarkers from 2 or more studies were represented in the pathway. Biomarkers that were not proteins or RNA were excluded from pathway analysis, as they were not compatible with pathway analysis.

## Results

### Study characteristics

The search yielded 11980 studies for review before duplicates were removed (Figure 1). Eleven studies^23–33^ reporting data from 1546 patients and eleven animals (macaques and mice) were eligible for the systematic review (See Table S1), as there was a precedent for studies combining both human and animal models contributing to cardiovascular disease management^34^. Of these, nine studies^25–33^ were conducted in humans, one study in animals (macaques)^24^, and one study in both animal (mice) and human cohorts (Table 1)^23^. All studies were published in English, and the publication years ranged from 2011 to 2025. Seven studies^25–27, 29, 31–33^ reported risk factors, of which three^25, 28, 31^ reported factors separately for cases and controls. Five studies^25, 28, 31–33^ reported the TOAST^35^ classification of stroke aetiologies, two studies^26, 27^ only included patients with large vessel occlusion, and one study^24^ occluded a large artery as part of experimental design in animals. Seven^25–28, 31–33^ studies reported on the use of reperfusion therapy, and one study^24^ reperfused all animals as part of experimental design.

**Figure 1.**
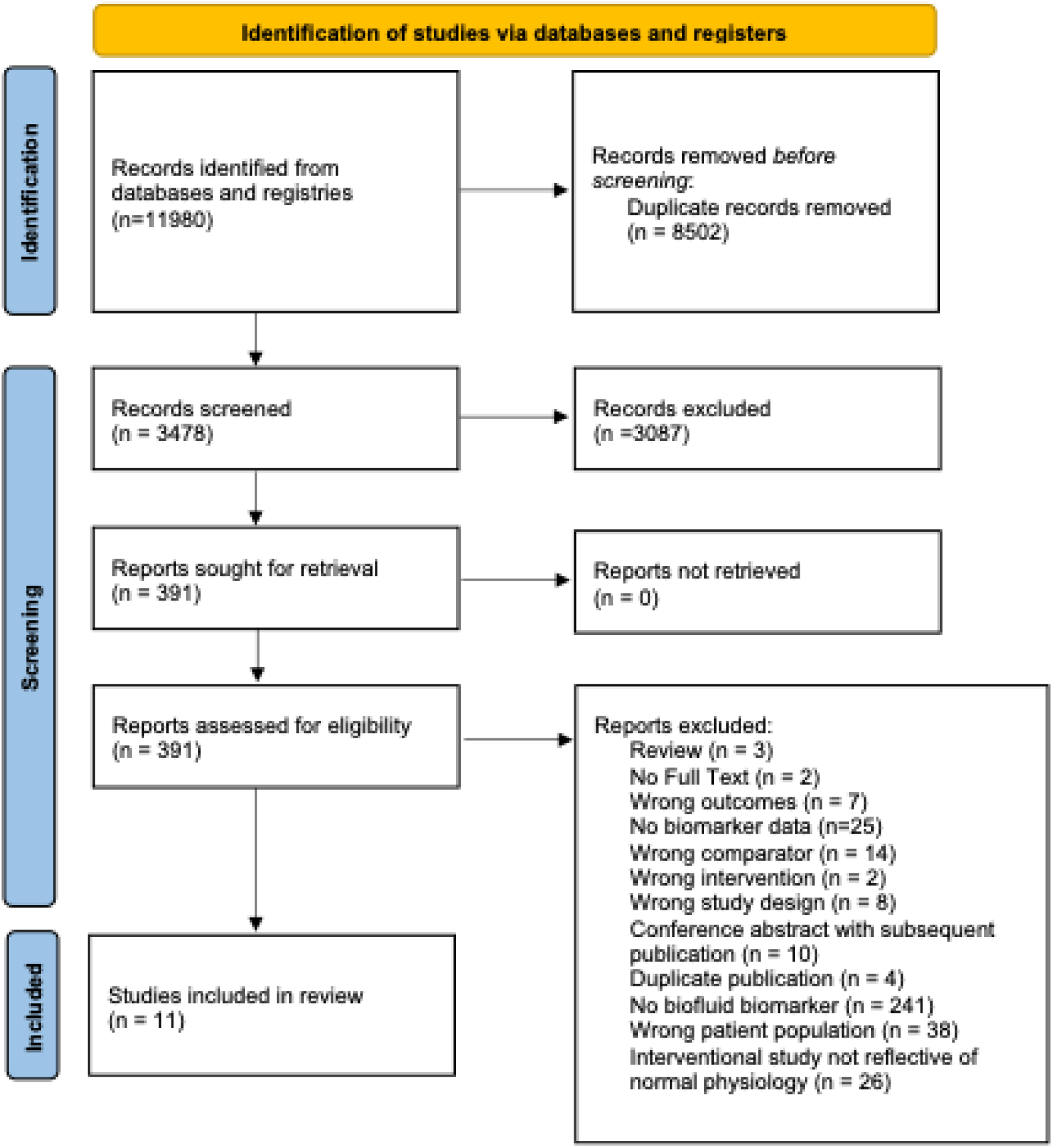
PRISMA diagram of studies included in the systematic review.

**Table 1:**
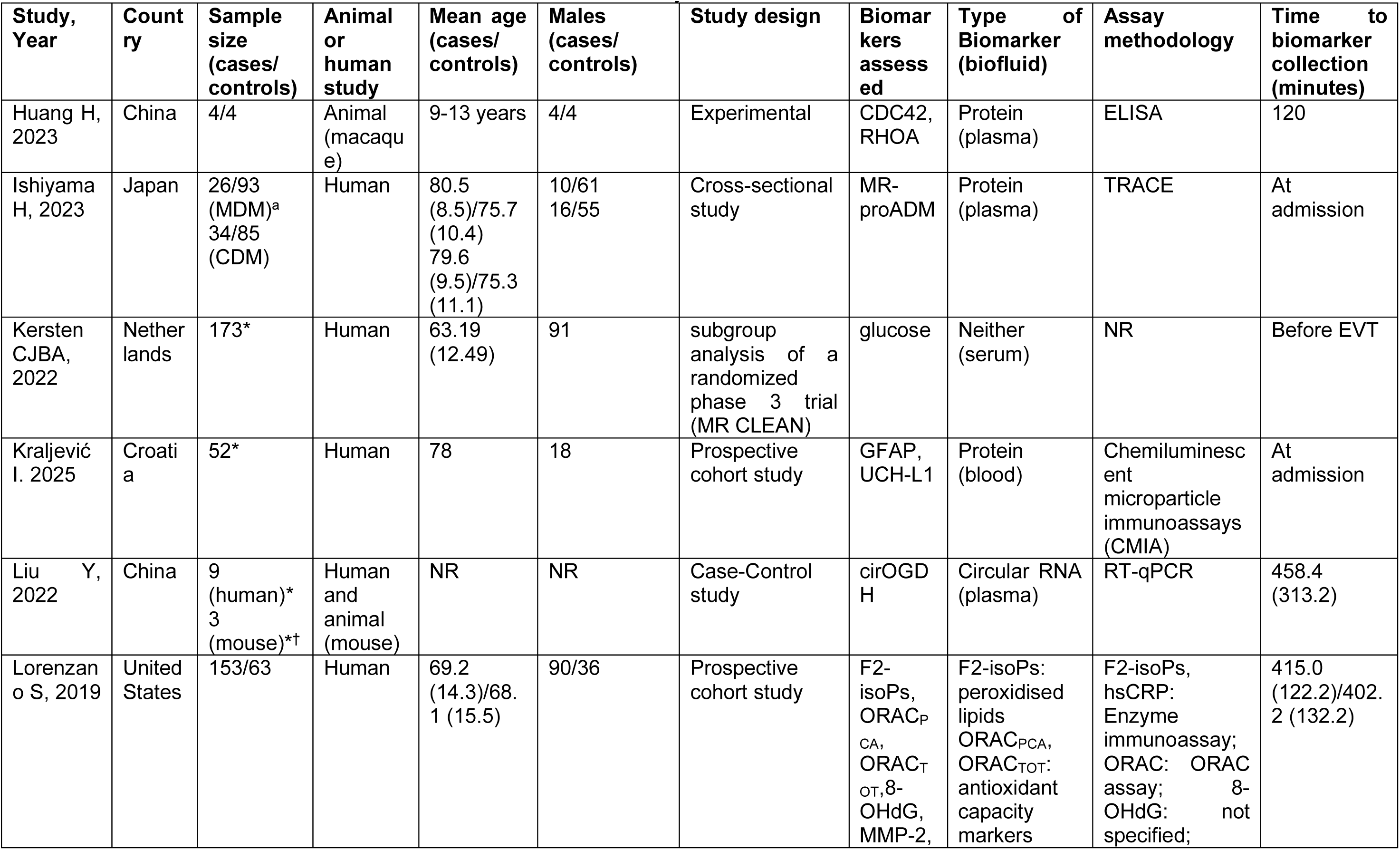
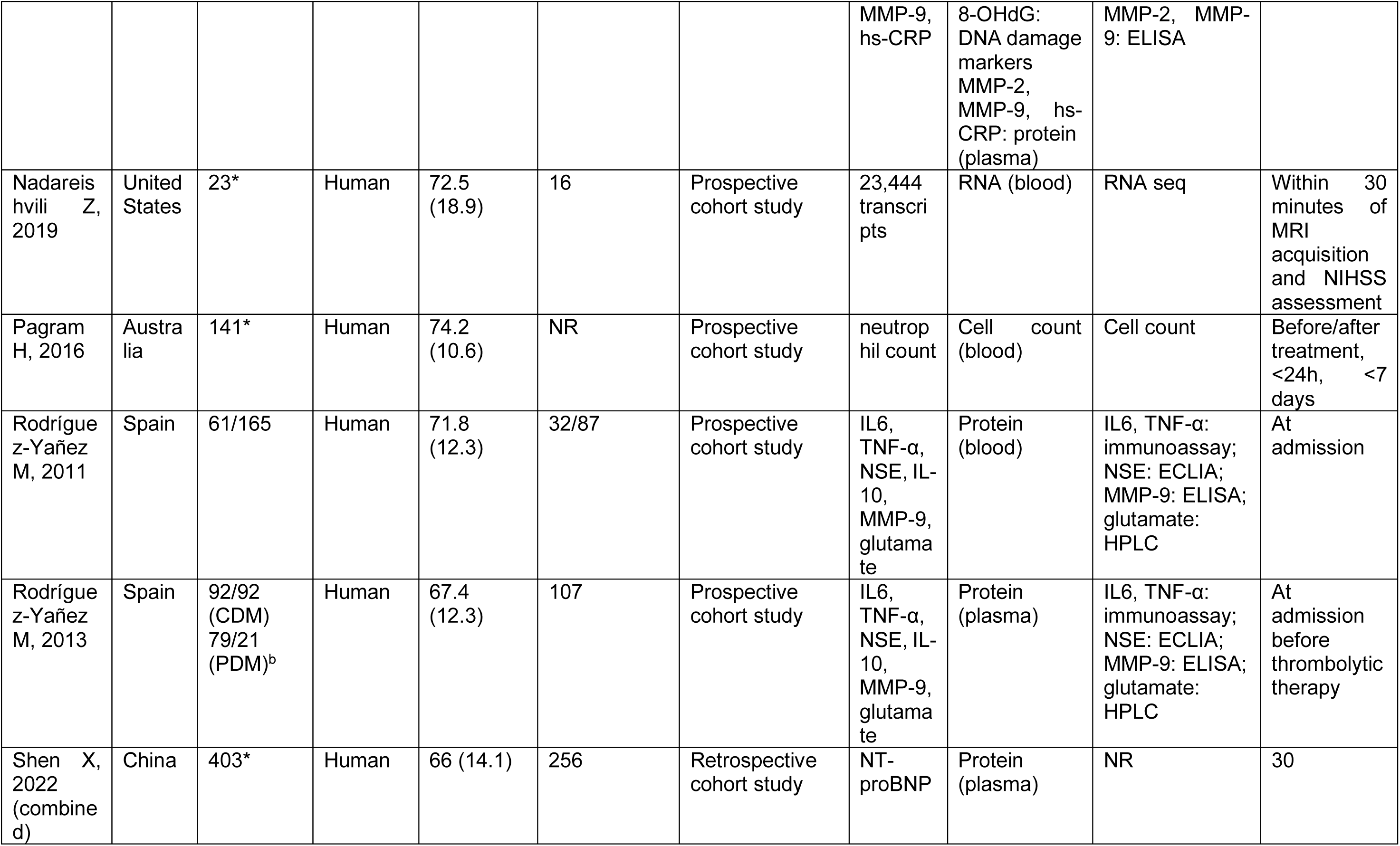
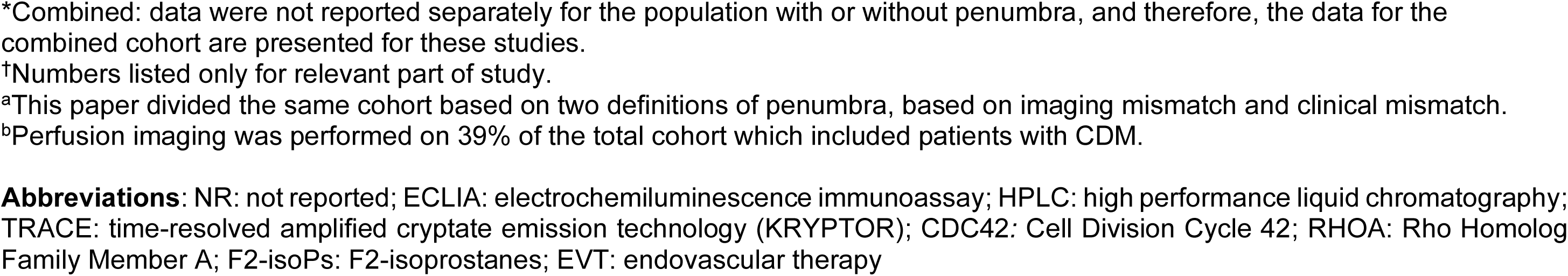
Baseline characteristics of the studies included in the systematic review.

| Study, Year | Country | Sample size (cases/controls) | Animal or human study | Mean age (cases/controls) | Males (cases/controls) | Study design | Biomarkers assessed | Type of Biomarker (biofluid) | Assay methodology | Time to biomarker collection (minutes) |
| --- | --- | --- | --- | --- | --- | --- | --- | --- | --- | --- |
| Huang H, 2023 | China | 4/4 | Animal (macaque) | 9-13 years | 4/4 | Experimental | CDC42, RHOA | Protein (plasma) | ELISA | 120 |
| Ishiyama H, 2023 | Japan | 26/93 (MDM)<sup>a</sup><br>34/85 (CDM) | Human | 80.5 (8.5)/75.7 (10.4)<br>79.6 (9.5)/75.3 (11.1) | 10/61<br>16/55 | Cross-sectional study | MR-proADM | Protein (plasma) | TRACE | At admission |
| Kersten CJBA, 2022 | Netherlands | 173* | Human | 63.19 (12.49) | 91 | subgroup analysis of a randomized phase 3 trial (MR CLEAN) | glucose | Neither (serum) | NR | Before EVT |
| Kraljević I. 2025 | Croatia | 52* | Human | 78 | 18 | Prospective cohort study | GFAP, UCH-L1 | Protein (blood) | Chemiluminescent microparticle immunoassays (CMIA) | At admission |
| Liu Y, 2022 | China | 9 (human)*<br>3 (mouse)*† | Human and animal (mouse) | NR | NR | Case-Control study | cirOGDH | Circular RNA (plasma) | RT-qPCR | 458.4 (313.2) |
| Lorenzano S, 2019 | United States | 153/63 | Human | 69.2 (14.3)/68.1 (15.5) | 90/36 | Prospective cohort study | F2-isoPs, ORAC<sub>PCA</sub>, ORAC<sub>TOT</sub>, 8-OHdG, MMP-2, | F2-isoPs: peroxidised lipids<br>ORAC<sub>PCA</sub>, ORAC<sub>TOT</sub>: antioxidant capacity markers | F2-isoPs, hsCRP: Enzyme immunoassay; ORAC: ORAC assay; 8-OHdG: not specified; | 415.0 (122.2)/402.2 (132.2) |
|  |  |  |  |  |  |  | MMP-9, hs-CRP | 8-OHdG: DNA damage markers<br>MMP-2, MMP-9, hs-CRP: protein (plasma) | MMP-2, MMP-9: ELISA |  |
| Nadareishvili Z, 2019 | United States | 23* | Human | 72.5 (18.9) | 16 | Prospective cohort study | 23,444 transcripts | RNA (blood) | RNA seq | Within 30 minutes of MRI acquisition and NIHSS assessment |
| Pagram H, 2016 | Australia | 141* | Human | 74.2 (10.6) | NR | Prospective cohort study | neutrophil count | Cell count (blood) | Cell count | Before/after treatment, <24h, <7 days |
| Rodríguez-Yañez M, 2011 | Spain | 61/165 | Human | 71.8 (12.3) | 32/87 | Prospective cohort study | IL6, TNF-α, NSE, IL-10, MMP-9, glutamate | Protein (blood) | IL6, TNF-α: immunoassay; NSE: ECLIA; MMP-9: ELISA; glutamate: HPLC | At admission |
| Rodríguez-Yañez M, 2013 | Spain | 92/92 (CDM)<br>79/21 (PDM)<sup>b</sup> | Human | 67.4 (12.3) | 107 | Prospective cohort study | IL6, TNF-α, NSE, IL-10, MMP-9, glutamate | Protein (plasma) | IL6, TNF-α: immunoassay; NSE: ECLIA; MMP-9: ELISA; glutamate: HPLC | At admission before thrombolytic therapy |
| Shen X, 2022 (combined) | China | 403* | Human | 66 (14.1) | 256 | Retrospective cohort study | NT-proBNP | Protein (plasma) | NR | 30 |

\*Combined: data were not reported separately for the population with or without penumbra, and therefore, the data for the combined cohort are presented for these studies.
†Numbers listed only for relevant part of study.
<sup>a</sup>This paper divided the same cohort based on two definitions of penumbra, based on imaging mismatch and clinical mismatch.
<sup>b</sup>Perfusion imaging was performed on 39% of the total cohort which included patients with CDM.
**Abbreviations:** NR: not reported; ECLIA: electrochemiluminescence immunoassay; HPLC: high performance liquid chromatography; TRACE: time-resolved amplified cryptate emission technology (KRYPTOR); CDC42: Cell Division Cycle 42; RHOA: Rho Homolog Family Member A; F2-isoPs: F2-isoprostanes; EVT: endovascular therapy

**Table 2.** Risk of Bias Assessment of Studies included in the Systematic Review.

| Study | Risk of bias |  |  |  | Applicability concerns |  |  |
| --- | --- | --- | --- | --- | --- | --- | --- |
|  | P | I | R | FT | P | I | R |
| Huang, 2023 | X | ✓ | ? | ✓ | X | ✓ | X |
| Ishiyama, 2023 | ✓ | ✓ | ? | ✓ | ✓ | ✓ | ✓ |
| Kersten, 2022 | X | X | ? | ✓ | ✓ | ✓ | ✓ |
| Kraljević I, 2025 | X | ✓ | ✓ | ✓ | ✓ | ✓ | ✓ |
| Liu, 2022 | X | ✓ | ? | ? | ✓ | ✓ | ✓ |
| Lorenzano, 2019 | ✓ | ✓ | ✓ | ✓ | ✓ | ✓ | ✓ |
| Nadareishvili, 2019 | X | ✓ | ? | ✓ | ✓ | ✓ | ✓ |
| Pagram, 2016 | ✓ | ✓ | ? | X | ✓ | ✓ | ✓ |
| Rodriguez-Yanez, 2011 | X | ✓ | ✓ | X | ✓ | ✓ | X |
| Rodriguez-Yanez, 2013 | X | X | ✓ | X | ✓ | ✓ | ✓ |
| Shen, 2022 | ✓ | ✓ | ✓ | ✓ | ✓ | ✓ | ✓ |

P = patient selection; I = index test; R = reference standard; FT = flow and timing. ✓ indicates low risk; X indicates high risk; ? indicates unclear risk.

### Risk of Bias

Two studies^28, 33^ had a low risk of bias, whereas nine studies^23–27, 29–32^ had some concerns or a high risk of bias. Unclear blinding status of reviewers of the reference test to the results of the index test was the most common reason for unclear risk of bias in the reference test domain. Nine studies^23, 25–30, 32, 33^ did not have concerns regarding applicability, whereas two studies^24, 31^ had concerns regarding applicability in the reference test domain. One study^31^ was marked as having concerns regarding applicability in patient selection since penumbra was identified solely using clinical criteria, which was not congruent with the current inclusion criteria for thrombectomy^15^.

### Imaging characteristics

Imaging modalities used in the studies to identify or quantify penumbra included MRI^23–25, 28, 29, 31, 32^ (seven studies; Table S2) and CT^26, 27, 30, 33^ (four studies). The presence of penumbra was reported as a dichotomous variable in five studies^24, 25, 28, 31, 32^, whereas six studies^23, 26, 27, 29, 30, 33^ reported penumbra volume as a continuous variable. Mismatch used for penumbra identification varied widely, including DWI-FLAIR^23, 24^, MRI perfusion-diffusion^28, 29, 32^, CTP^26, 27, 33^, CTP-DWI^30^, DWI-clinical^25, 31^ and MRA-DWI^25^ mismatches (Table S2). Of the 4 studies reporting thresholds for the identification of penumbra, two studies^28, 32^ used >20% mismatch ratio as the primary cutoff for penumbra presence, of which 1 used PWI-DWI ≥ 10 cm³, PWI ≥ 10 cm³ as an additional “strict” criterion^28^. One study^25^ used the MRA-DWI mismatch criteria^36^ as well as the clinical diffusion mismatch criteria (NIHSS ≥8 and DWI lesion ≤25mL)^37^ for the same cohort, and one study^31^ used the clinical diffusion mismatch criteria. One study^24^ used visual inspection to assess for the presence of DWI-FLAIR mismatch. Additional imaging definitions and characteristics are reported in Table S2. This heterogeneity in penumbra definition likely contributed to variability in biomarker associations and limits cross-study comparability, underscoring the need for standardized imaging criteria in future biomarker validation studies.

### Biomarker Findings

The studies reported results for 53 biomarkers in humans, which included 34 transcripts, 11 proteins, one amino acid, two markers of antioxidant capacity, one marker of DNA damage, one circular RNA, and one peroxidised lipid, as well as neutrophil count and glucose (Table 1). One human biomarker was independently verified in mice (circOGDH; gene expression), and two biomarkers (CDC42 and RHOA; proteins) were reported only in animals.

All biomarkers were measured in blood or plasma, collected at different timepoints as reported in Table 1. Data stratified on clinical-diffusion mismatch was used for Rodriguez-Yañez 2013^32^ to avoid selection bias restricting to patients receiving perfusion imaging. Of these, sufficient data were available to calculate SMDs between groups with and without penumbra for 14 biomarkers across four^25, 28, 31, 32^studies (Table 3). Meta-analyses were performed on six biomarkers which were assessed in more than one study (Figure 2). Significant associations were observed for two biomarkers: interleukin-10 (IL-10; SMD 2.40, 95% CI 1.37 to 3.42), which was elevated in the penumbra-positive group, and interleukin-6 (IL-6; SMD -0.93; 95% CI -1.80 to -0.06), which was reduced in the penumbra-positive group compared with patients without penumbrae. MMP-9, NSE, TNF-α, and glutamate were not significantly associated with penumbrae. These findings should be interpreted cautiously due to high heterogeneity across contributing studies.

**Figure 2.**
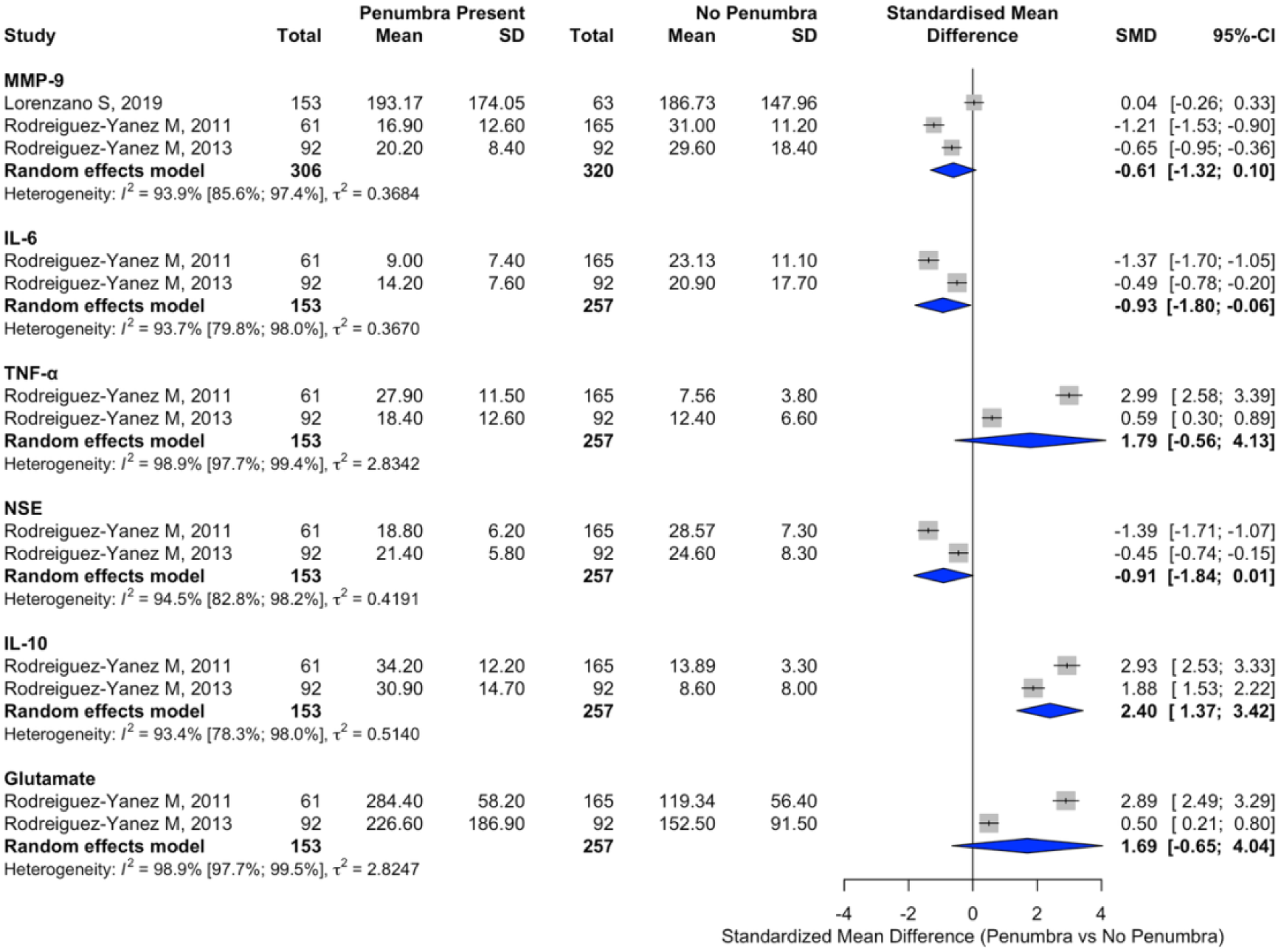
Forest Plot Representing Pooled Synthesis for Biomarkers Included in the Meta-Analysis.

**Table 3.** Biomarkers with Calculated Standardised Mean Differences between Patients with and without Ischaemic Penumbrae.

| **Biomarker** | **No of cases** | **No of controls** | **No of studies** | **SMD\* (95% CI); I<sup>2</sup>** | **p-value** | **Study reference** |
| --- | --- | --- | --- | --- | --- | --- |
| MR-proADM (MDM) | 26 | 93 | 1 | 0.97 (0.52 to 1.42) | <0.001 | 25 |
| MR-proADM (CDM) | 34 | 85 | 1 | 0.88 (0.47 to 1.30) | <0.001 | 25 |
| IL-10 | 153 | 257 | 2 | 2.40 (1.37 to 3.42); 93.4 | <0.01 | 31, 32 |
| IL-6 | 153 | 257 | 2 | -0.93 (-1.80 to -0.06); 93.7 | 0.04 | 31, 32 |
| ORAC<sub>PCA</sub> | 153 | 63 | 1 | 0.31 (0.01 to 0.60) | 0.04 | 28 |
| NSE | 153 | 257 | 2 | -0.91 (-1.84 to 0.01); 94.5 | 0.05 | 31, 32 |
| MMP-9 | 306 | 320 | 3 | -0.61 (-1.32 to 0.10); 93.9 | 0.09 | 28, 31, 32 |
| TNF-α | 153 | 257 | 2 | 1.79 (-0.56 to 4.13); 98.9 | 0.14 | 31, 32 |
| glutamate | 153 | 257 | 2 | 1.69 (-0.65 to 4.04); 98.9 | 0.15 | 31, 32 |
| F2-isoPs | 153 | 63 | 1 | 0.18 (-0.10 to 0.48) | 0.21 | 28 |
| ORAC<sub>TOT</sub> | 153 | 63 | 1 | -0.14 (-0.43 to 0.15) | 0.35 | 28 |
| 8-OHdG | 153 | 63 | 1 | -0.14 (-0.44 to 0.15) | 0.34 | 28 |
| 8-OHdG (adjusted for creatinine) | 153 | 63 | 1 | -0.11 (-0.40 to 0.18) | 0.46 | 28 |
| MMP-2 | 153 | 63 | 1 | 0.11 (-0.17 to 0.41) | 0.44 | 28 |
| hs-CRP | 153 | 63 | 1 | 0.11 (-0.19 to 0.39) | 0.50 | 28 |

\*SMD: Standardised Mean Difference; MDM: MRA-Diffusion Mismatch, CDM: Clinical-Diffusion Mismatch

Amongst biomarkers for which a meta-analysis was not possible, 38 biomarkers demonstrated significant SMDs with presence of penumbra or correlation with penumbra volume in humans. Radical absorbance capacity after perchloric acid treatment (ORAC_PCA_; SMD 0.31; 95% CI 0.01 to 0.60) and mid-regional pro-adrenomedullin (MR-proADM; SMD 0.97; 95% CI 0.52 to 1.42 for MDM, see Table 1 for CDM) significantly associated with presence of a penumbra (Table 3), and significant correlations were observed for 35 RNA biomarkers and NT-proBNP with penumbra volume (Table 4). Correlation of circOGDH with penumbra volume was independently verified in mice (Table 4). MMP-2, 8-OHdG, hs-CRP, total radical absorbance capacity (ORAC_TOT_), and F2-isoPs did not show significant SMDs. Neutrophil count, GFAP, and UCH-L1 were not significantly correlated with penumbra volume.

**Table 4.** Correlation of Biomarkers with Penumbra Volume.

| Biomarker | No of patients (combined) | No of studies | Correlation coefficient (p-value) | Study reference |
| --- | --- | --- | --- | --- |
| *STK26* | 23 | 1 | 0.58 (0.003) | 29 |
| *MGA* | 23 | 1 | 0.58 (0.004) | 29 |
| *UBA2* | 23 | 1 | 0.57 (0.004) | 29 |
| *PICALM* | 23 | 1 | 0.57 (0.004) | 29 |
| *CDK11B* | 23 | 1 | 0.57 (0.004) | 29 |
| *UXT* | 23 | 1 | 0.56 (0.005) | 29 |
| *S100A6* | 23 | 1 | 0.55 (0.006) | 29 |
| *TES* | 23 | 1 | 0.55 (0.007) | 29 |
| *GABARAPL2* | 23 | 1 | 0.54 (0.008) | 29 |
| *HDAC1* | 23 | 1 | 0.53 (0.008) | 29 |
| *CORO1A* | 23 | 1 | 0.53 (0.009) | 29 |
| *SLC38A2* | 23 | 1 | 0.53 (0.009) | 29 |
| *POLG* | 23 | 1 | 0.52 (0.011) | 29 |
| *CNN2* | 23 | 1 | 0.52 (0.011) | 29 |
| *EBP* | 23 | 1 | 0.52 (0.011) | 29 |
| *GLTP* | 23 | 1 | 0.49 (0.018) | 29 |
| *RPS2* | 23 | 1 | 0.47 (0.022) | 29 |
| *NUP98* | 23 | 1 | -0.71 (<0.001) | 29 |
| *SEMA4D* | 23 | 1 | -0.64 (0.001) | 29 |
| *HCAR3* | 23 | 1 | -0.61 (0.002) | 29 |
| *IL1B* | 23 | 1 | -0.59 (0.003) | 29 |
| *SENP6* | 23 | 1 | -0.57 (0.005) | 29 |
| *PF4* | 23 | 1 | -0.56 (0.005) | 29 |
| *LINC-PINT* | 23 | 1 | -0.54 (0.007) | 29 |
| *HCAR2* | 23 | 1 | -0.52 (0.011) | 29 |
| *GP9* | 23 | 1 | -0.51 (0.012) | 29 |
| *DYNC1LI1* | 23 | 1 | -0.51 (0.013) | 29 |
| *FFAR2* | 23 | 1 | -0.49 (0.017) | 29 |
| *ZNF117* | 23 | 1 | -0.50 (0.017) | 29 |
| *PUM2* | 23 | 1 | -0.49 (0.018) | 29 |
| *RAB27B* | 23 | 1 | -0.48 (0.021) | 29 |
| *KCNJ2* | 23 | 1 | -0.48 (0.021) | 29 |
| *TCEA1* | 23 | 1 | -0.48 (0.021) | 29 |
| *GNB2* | 23 | 1 | -0.47 (0.024) | 29 |
| NT-proBNP | 403 | 1 | 0.199 (<0.001) | 33 |
| GFAP | 51 | 1 | 0.112 (0.428) | 27 |
| UCH-L1 | 52 | 1 | -0.053 (0.708) | 27 |
| circOGDH | 9 | 1 | 0.962 (0.002)* | 23 |
| Neutrophil count | 141 | 1 | NR; $R^2 = 0.217$ (0.212) | 30 |

\*Mouse: $r = 0.847$ (0.001), $n = 3$

Although there were insufficient data to calculate SMDs or assess correlation, the core-penumbra ratio was lower in hyperglycaemic patients than in normoglycaemic patients (p = 0.02)^26^, although penumbra volume did not differ significantly (p = 0.06). In non-human primates, CDC42 (cell division cycle 42) expression was reported to be lower in the penumbra-negative group (p < 0.05)^24^, but there was insufficient information to calculate SMDs.

### Biological Pathways Underlying Penumbra Biomarkers

PPI network analysis was subsequently performed on 38 protein or RNA biomarkers identified as having a significant association with penumbrae based on SMDs or correlation coefficients in either humans or animals. LINC-PINT, circOGDH, and ORAC_PCA_ were excluded because they do not have protein products which lend themselves to analysis.

The interaction network consisted of 18 highly connected biomarkers (nodes) having 28 interactions (edges). Our analysis demonstrated that *IL1B* encoding interleukin-1β (IL-1β) had the highest number of interactions with other proteins (10 interactions), followed by IL-6 with 9 interactions, IL-10 with 5 interactions, histone deacetylase 1 (HDAC1) and hydroxycarboxylic acid receptor 2 (HCAR2) with 4 interactions, and free fatty acid receptor 2 (FFAR2), platelet factor 4 (PF4, a.k.a. CXCL4), and CDC42 each with 3 interactions (Figure 3A). IL-6/IL-10, IL-1β/IL-6 and IL-1β/IL-10 exhibited the strongest interactions, with functional interaction probabilities of 0.999, 0.997, 0.996, respectively.

**Figure 3.**
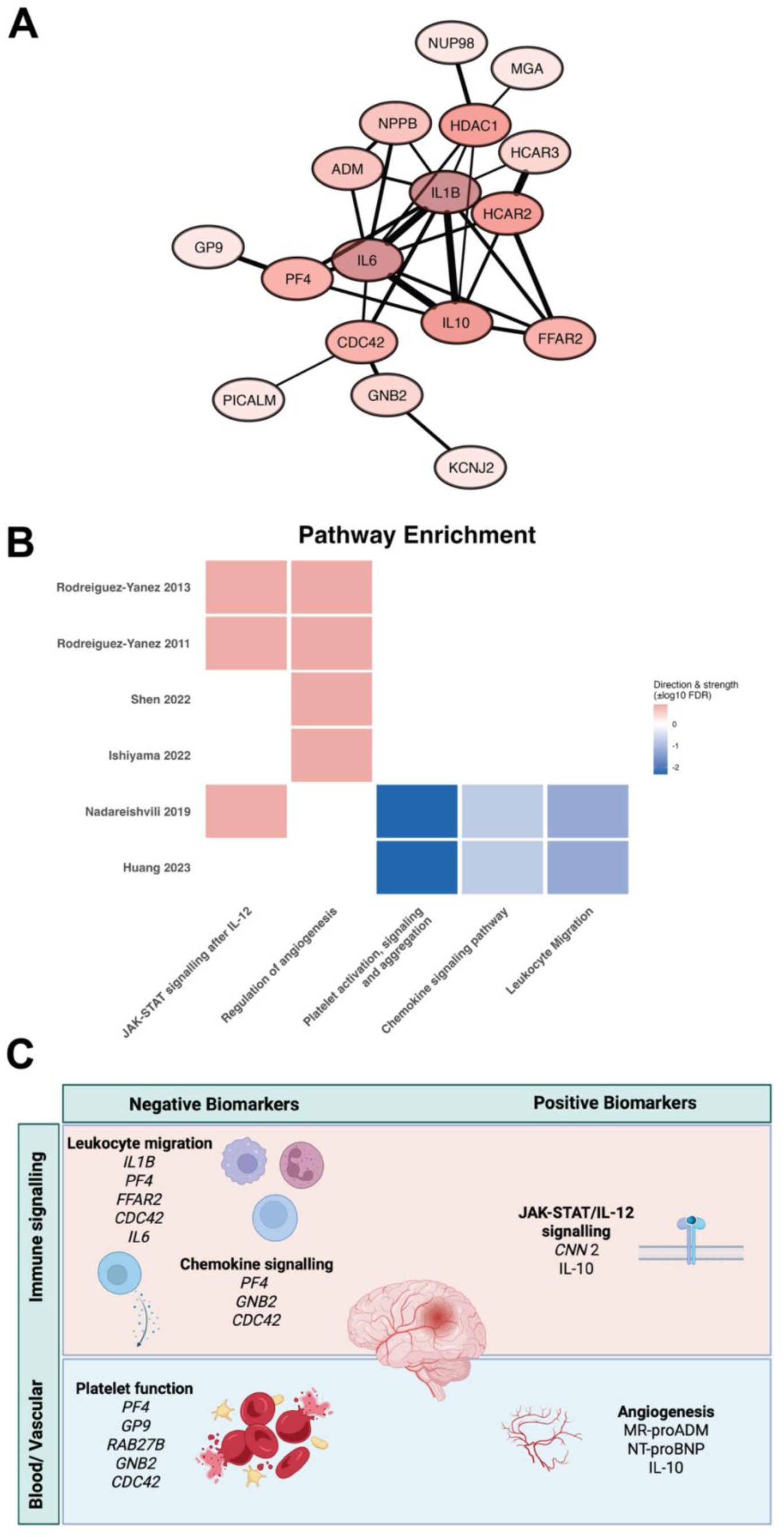
Gene Pathways Associated with the Presence of Penumbrae. **A**. Protein-protein interaction network of significant biomarkers. Node colours represent the number of interactions, with the darker colour representing more interactions, whereas edges connecting nodes represent the strength of interactions, with more thickness representing stronger interactions among proteins. **B**. Pathway enrichment heatmap demonstrating upregulated (red) and downregulated (blue) pathways, respectively. **C**. Schema demonstrating gene pathways associated with identified biomarkers; *NPPB* encodes BNP.

Pathways related to angiogenesis and interleukin-12 signalling were enriched among biomarkers positively associated with penumbra presence or volume (Figure 3B, C). In contrast, pathways involving leukocyte migration, chemokine signalling, and platelet activation were enriched among biomarkers inversely associated with presence of penumbra (Figure 3B, C).

## Discussion

This systematic review analysed 55 biomarkers from 1546 patients and 11 animals and identified two human biomarkers (IL-10, IL-6) that were significantly associated with the presence of a penumbra in the meta-analysis, as well as 38 biomarkers with reported significance in individual studies in humans (MR-proADM, ORAC_PCA_, circOGDH, NT-proBNP, etc.). *Cdc42* expression was reported to be significantly associated with penumbra presence in nonhuman primates, but this association could not be verified using SMD analysis. Integrative analysis was limited because results were presented in different ways, including odds ratios^25, 28, 31, 32^, Pearson’s coefficient^26, 27, 29^, Spearman’s rho^33^, r^2^ values^30^, and presence of a significant difference^24^. Limited overlap amongst the biomarkers being tested also hindered integration. Regardless, common themes emerged in the subsequent gene pathway analysis, linking pathways related to the immune system, haemostasis, and angiogenesis to the presence of penumbrae. Despite limited current evidence on the predictive capacity of circulating biomarkers of ischaemic penumbra, the biomarkers highlighted in the current review may have utility beyond their potential roles in clinical decision-making, as they can provide insights into targets for post-stroke neuroprotection.

The convergence of identified biomarkers on immune modulation and haemostatic signalling suggests that penumbra biology reflects a dynamic balance between tissue-protective and tissue-injurious processes. Biomarkers such as IL-10 and MR-proADM may reflect compensatory vasodilatory and anti-inflammatory responses that preserve microvascular perfusion, whereas IL-1β, IL-6, platelet-derived mediators, and neutrophil-associated genes may signal transition toward irreversible infarction.

Biomarkers positively associated with penumbra included MR-proADM (adrenomedullin) and IL-10, both of which have vasodilatory and anti-inflammatory functions^38, 39^. This finding, if confirmed in future investigations, may have translational implications, as the presence of these two biomarkers may benefit the ischaemic brain. For example, IL-10 was upregulated in infiltrating T cells and macrophages in a mouse model of transient MCA occlusion and exerted a neuroprotective effect by suppressing IL-17 production, a pro-inflammatory cytokine, and by preventing infarct growth. In humans, lower levels of IL-10 production were linked to a higher incidence of stroke^40^. Furthermore, adrenomedullin promotes angiogenesis by upregulating VEGF^41^, and IL-10 overcomes pro-inflammatory IFN-γ- mediated suppression of VEGF^42^, thereby promoting angiogenesis. Together, modulation of type 1 immune responses and promotion of cerebral angiogenesis are likely to act in tandem to extend the viability of penumbra tissue and thus serve as biomarkers of bioprocesses that maintain salvageable tissue.

Markers associated with leukocyte migration and chemokine signalling, as well as regulators of platelet function, were negative biomarkers of ischaemic penumbra. IL-1β is known to trigger apoptosis of injured cells^43^, and both IL-1β and GNB2 promote neutrophil migration^43, 44^, which can lead to oxidative damage. Similarly, IL-6 expression in CSF^45^ and blood^46^ correlates with lesion size and worse functional outcomes, likely due to production of inflammatory mediators including reactive oxygen species and upregulating leukocyte adhesion molecules^47^. Markers associated with platelet activation, such as PF4 (a.k.a. CXCL4), which is a powerful driver of platelet aggregation,^48^ were also negatively associated with the presence of ischaemic penumbra. Taken together, these negative biomarkers suggest that pro-inflammatory and pro-coagulative pathways can promote secondary neurological injury via apoptosis and hypoxia, which reduces penumbra salvageability.

The main purpose of this review was to identify previously reported biomarkers that can help select patients with penumbra. The findings suggest that although these biomarkers show promise, they need to be tested in rigorously designed discovery and validation studies using methods acceptable to regulatory bodies, e.g., the US FDA. If prospectively validated, rapid multiplex biomarker panels could complement clinical stroke scales in prehospital triage, particularly in regions without immediate access to advanced imaging. They could be incorporated into prehospital point-of-care (POC) tests, analogous to capillary glucose or POC troponin and lactate, which are currently being explored for acute coronary syndrome and sepsis, respectively^49^. Penumbra biomarker tests could enhance the accuracy of prehospital large-vessel occlusion scores (which currently have a sensitivity of 47-73%^50^) to guide direct transport to a thrombectomy centre (Figure 4). In addition, detection of penumbra biomarkers in patients with resolved ischaemic stroke symptoms would further enhance our confidence in distinguishing a transient ischaemic attack from TIA mimics such as seizures.

**Figure 4.**
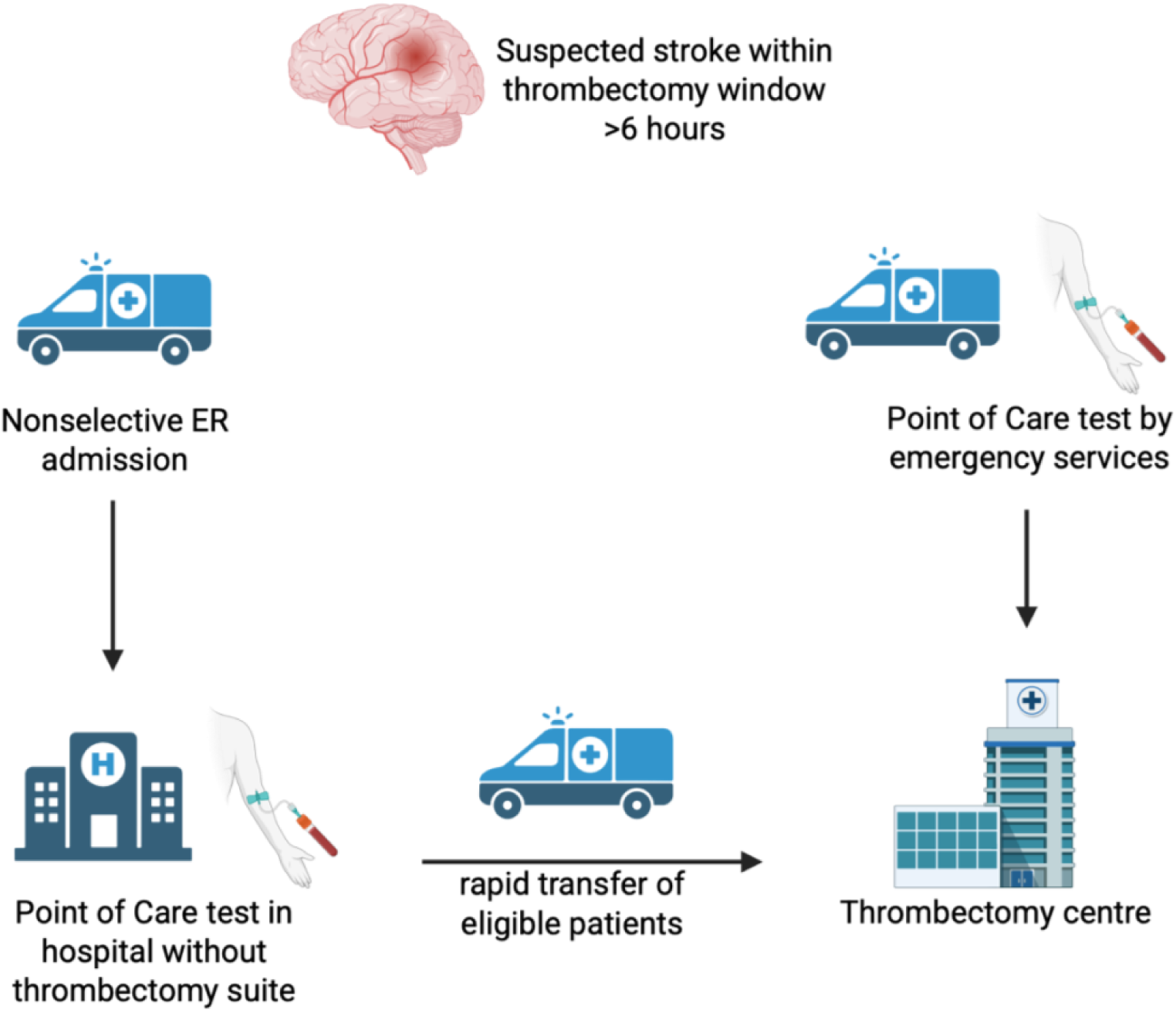
Schema of Possible Biomarker-Based Stratification of Stroke Admissions for Mechanical Thrombectomy. Figure created with BioRender.

We acknowledge several limitations of the study. The sample sizes of many of the included studies were small, especially those using animal models, limiting statistical power. In addition, we were unable to conduct a meta-analysis on the majority of biomarkers due to minimal overlap between studies, and the number of pooled studies was limited even where a meta-analysis was possible. Inconsistent reporting of biomarker levels and penumbra definitions by the included studies hindered standardised comparisons. Variability in the timing of biomarker measurements could also contribute to length-time bias, and differences in biomarker levels between groups could be confounded by stroke severity. Furthermore, most included studies had a risk of bias. We were unable to assess publication bias because the number of pooled studies per biomarker was small.

In summary, this systematic review identified biomarkers of ischaemic penumbra and highlighted the central role of pathways regulating the inflammatory response and cerebral perfusion in predicting the presence of penumbra. Future research should focus on validating the roles of the IL-10, IL-6, and IL-1β pathways as core markers of penumbral immune signatures, and on investigating various regulators of platelet activity as candidates for novel biomarkers. Markers of oxidative stress also appear to be promising candidates for biomarkers but require complex sample processing and specialised equipment to be measured. Future work should prioritise standardized imaging definitions, prospective multi-centre validation cohorts, and integration of biomarker panels with clinical and imaging data. Only through rigorous validation can biofluid biomarkers transition from exploratory signals to clinically actionable tools for penumbra-guided stroke care.

## Data Availability

All data can be found in the original publications.

## References

1. Demeestere J, Wouters A, Christensen S, Lemmens R, Lansberg MG. Review of Perfusion Imaging in Acute Ischemic Stroke: From Time to Tissue. Stroke 2020;51:1017–1024.

2. Emberson J, Lees KR, Lyden P, et al. Effect of treatment delay, age, and stroke severity on the effects of intravenous thrombolysis with alteplase for acute ischaemic stroke: a meta-analysis of individual patient data from randomised trials. Lancet 2014;384:1929–1935.

3. Hacke W, Kaste M, Bluhmki E, et al. Thrombolysis with alteplase 3 to 4.5 hours after acute ischemic stroke. N Engl J Med 2008;359:1317–1329.

4. Thomalla G, Simonsen CZ, Boutitie F, et al. MRI-Guided Thrombolysis for Stroke with Unknown Time of Onset. New England Journal of Medicine 2018;379:611–622.

5. Ringleb P, Bendszus M, Bluhmki E, et al. Extending the time window for intravenous thrombolysis in acute ischemic stroke using magnetic resonance imaging-based patient selection. International Journal of Stroke 2019;14:483–490.

6. Ma H, Campbell BCV, Parsons MW, et al. Thrombolysis Guided by Perfusion Imaging up to 9 Hours after Onset of Stroke. New England Journal of Medicine 2019;380:1795–1803.

7. Koga M, Yamamoto H, Inoue M, et al. Thrombolysis With Alteplase at 0.6 mg/kg for Stroke With Unknown Time of Onset. Stroke 2020;51:1530–1538.

8. Wang L, Dai Y-J, Cui Y, et al. Intravenous Tenecteplase for Acute Ischemic Stroke Within 4.5–24 Hours of Onset (ROSE-TNK): A Phase 2, Randomized, Multicenter Study. J Stroke 2023;25:371–377.

9. Günkan A, Ferreira MY, Vilardo M, et al. Thrombolysis for Ischemic Stroke Beyond the 4.5-Hour Window: A Meta-Analysis of Randomized Clinical Trials. Stroke 2025;56:580–590.

10. Mishra NK, Albers GW, Christensen S, et al. Comparison of magnetic resonance imaging mismatch criteria to select patients for endovascular stroke therapy. Stroke 2014;45:1369–1374.

11. Mishra NK, Albers GW, Davis SM, et al. Mismatch-based delayed thrombolysis: a meta-analysis. Stroke 2010;41:e25–33.

12. Zachrison KS, Ganti L, Sharma D, et al. A survey of stroke-related capabilities among a sample of US community emergency departments. JACEP Open 2022;3.

13. Lee JY, Kim DY, Kim JY, et al. Perfusion Imaging-Based Triage for Acute Ischemic Stroke: Trends in Use and Impact on Clinical Outcomes. Stroke: Vascular and Interventional Neurology 2024;4:e001361.

14. Asdaghi N, Wang K, Gardener H, et al. Impact of Time to Treatment on Endovascular Thrombectomy Outcomes in the Early Versus Late Treatment Time Windows. Stroke 2023;54:733–742.

15. Powers WJ, Rabinstein AA, Ackerson T, et al. Guidelines for the Early Management of Patients With Acute Ischemic Stroke: 2019 Update to the 2018 Guidelines for the Early Management of Acute Ischemic Stroke: A Guideline for Healthcare Professionals From the American Heart Association/American Stroke Association. Stroke 2019;50:e344–e418.

16. Page MJ, McKenzie JE, Bossuyt PM, et al. The PRISMA 2020 statement: an updated guideline for reporting systematic reviews. BMJ 2021;372:n71.

17. Whiting PF, Rutjes AWS, Westwood ME, et al. QUADAS-2: A Revised Tool for the Quality Assessment of Diagnostic Accuracy Studies. Annals of Internal Medicine 2011;155:529–536.

18. Wan X, Wang W, Liu J, Tong T. Estimating the sample mean and standard deviation from the sample size, median, range and/or interquartile range. BMC Medical Research Methodology 2014;14:135.

19. Higgins JPT, Thompson SG, Deeks JJ, Altman DG. Measuring inconsistency in meta-analyses. BMJ 2003;327:557–560.

20. Gustavsen J, Pai S, Isserlin R, Demchak B, Pico A. RCy3: Network biology using Cytoscape from within R [version 3; peer review: 3 approved]. F1000Research 2019;8.

21. Szklarczyk D, Kirsch R, Koutrouli M, et al. The STRING database in 2023: protein–protein association networks and functional enrichment analyses for any sequenced genome of interest. Nucleic Acids Research 2022;51:D638 –D646.

22. Kuleshov MV, Jones MR, Rouillard AD, et al. Enrichr: a comprehensive gene set enrichment analysis web server 2016 update. Nucleic Acids Research 2016;44:W90–W97.

23. Liu Y, Li Y, Zang J, et al. CircOGDH Is a Penumbra Biomarker and Therapeutic Target in Acute Ischemic Stroke. Circulation Research 2022;130:907–924.

24. Huang H, Wu S, Liang C, et al. CDC42 Might Be a Molecular Signature of DWI-FLAIR Mismatch in a Nonhuman Primate Stroke Model. Brain sciences 2023;13.

25. Ishiyama H, Tanaka T, Saito S, et al. Plasma mid-regional pro-adrenomedullin: A biomarker of the ischemic penumbra in hyperacute stroke. Brain pathology (Zurich, Switzerland) 2023;33:e13110.

26. Kersten C, Zandbergen AAM, Berkhemer OA, et al. Association of hyperglycemia and computed tomographic perfusion deficits in patients who underwent endovascular treatment for acute ischemic stroke caused by a proximal intracranial occlusion: a subgroup analysis of a randomized phase 3 trial (MR CLEAN). Journal of the neurological sciences 2022;440:120333.

27. Kraljević I, Marinović Guić M, Budimir Mršić D, et al. Can Serum GFAP and UCH-L1 Replace CT in Assessing Acute Ischemic Stroke Severity? Life [serial online] 2025;15:495. Available at: https://mdpi-res.com/d_attachment/life/life-15-00495/article_deploy/life-15-00495-v2.pdf?version=1742352277.

28. Lorenzano S, Rost NS, Khan M, et al. Early molecular oxidative stress biomarkers of ischemic penumbra in acute stroke. Neurology 2019;93:e1288–e1298.

29. Nadareishvili Z, Kelley D, Luby M, et al. Molecular signature of penumbra in acute ischemic stroke: a pilot transcriptomics study. Annals of clinical and translational neurology 2019;6:817–820.

30. Pagram H, Bivard A, Lincz LF, Levi C. Peripheral Immune Cell Counts and Advanced Imaging as Biomarkers of Stroke Outcome. Cerebrovascular diseases extra 2016;6:120–128.

31. Rodríguez-Yáñez M, Sobrino T, Arias S, et al. Early Biomarkers of Clinical–Diffusion Mismatch in Acute Ischemic Stroke. Stroke 2011;42:2813–2818.

32. Rodriguez-Yanez M, Castellanos M, Sobrino T, et al. Interleukin-10 facilitates the selection of patients for systemic thrombolysis. BMC Neurol 2013;13:7.

33. Shen X, Liao J, Jiang Y, et al. Elevated NT-proBNP levels are associated with CTP ischemic volume and 90-day functional outcomes in acute ischemic stroke: a retrospective cohort study. BMC cardiovascular disorders 2022;22:431.

34. Wira CR, Becker JU, Martin G, Donnino MW. Anti-arrhythmic and vasopressor medications for the treatment of ventricular fibrillation in severe hypothermia: A systematic review of the literature. Resuscitation 2008;78:21–29.

35. Adams HP, Bendixen BH, Kappelle LJ, et al. Classification of subtype of acute ischemic stroke. Definitions for use in a multicenter clinical trial. TOAST. Trial of Org 10172 in Acute Stroke Treatment. Stroke 1993;24:35–41.

36. Lansberg MG, Thijs VN, Bammer R, et al. The MRA-DWI Mismatch Identifies Patients With Stroke Who Are Likely to Benefit From Reperfusion. Stroke 2008;39:2491–2496.

37. Dávalos A, Blanco M, Pedraza S, et al. The clinical–DWI mismatch. Neurology 2004;62:2187–2192.

38. Sun J, Qian P, Kang Y, et al. Adrenomedullin 2 attenuates LPS-induced inflammation in microglia cells by receptor-mediated cAMP-PKA pathway. Neuropeptides 2021;85:102109.

39. Singh K, Misra DP. Interleukin-10: Role in arterial wall homeostasis and dampening of inflammation in Takayasu arteritis. International Journal of Rheumatic Diseases 2023;26:1663–1666.

40. van Exel E, Gussekloo J, de Craen AJM, Bootsma-van der Wiel A, Frölich M, Westendorp RGJ. Inflammation and Stroke. Stroke 2002;33:1135–1138.

41. Maki T, Ihara M, Fujita Y, et al. Angiogenic and Vasoprotective Effects of Adrenomedullin on Prevention of Cognitive Decline After Chronic Cerebral Hypoperfusion in Mice. Stroke 2011;42:1122–1128.

42. Wu W-K, Llewellyn OPC, Bates DO, Nicholson LB, Dick AD. IL-10 regulation of macrophage VEGF production is dependent on macrophage polarisation and hypoxia. Immunobiology 2010;215:796–803.

43. Zhu H, Hu S, Li Y, et al. Interleukins and Ischemic Stroke. Frontiers in Immunology 2022;Volume 13 - 2022.

44. Block H, Stadtmann A, Riad D, et al. Gnb isoforms control a signaling pathway comprising Rac1, Plcβ2, and Plcβ3 leading to LFA-1 activation and neutrophil arrest in vivo. Blood 2016;127:314–324.

45. Tarkowski E, Rosengren L, Blomstrand C, et al. Early Intrathecal Production of Interleukin -6 Predicts the Size of Brain Lesion in Stroke. Stroke 1995;26:1393–1398.

46. Smith CJ, Emsley HCA, Gavin CM, et al. Peak plasma interleukin-6 and other peripheral markers of inflammation in the first week of ischaemic stroke correlate with brain infarct volume, stroke severity and long-term outcome. BMC Neurology 2004;4:2.

47. Su J-H, Luo M-Y, Liang N-, et al. Interleukin-6: A Novel Target for Cardio-Cerebrovascular Diseases. Frontiers in Pharmacology 2021;Volume 12 - 2021.

48. Liu Z-Y, Sun M-X, Hua M-Q, et al. New perspectives on the induction and acceleration of immune-associated thrombosis by PF4 and VWF. Frontiers in Immunology 2023;Volume 14 - 2023.

49. Moore THM, Dawson S, Kirby K, et al. Point-of-care tests in the emergency medical services: a scoping review. Scandinavian Journal of Trauma, Resuscitation and Emergency Medicine 2025;33:18.

50. Smith EE, Kent DM, Bulsara KR, et al. Accuracy of Prediction Instruments for Diagnosing Large Vessel Occlusion in Individuals With Suspected Stroke: A Systematic Review for the 2018 Guidelines for the Early Management of Patients With Acute Ischemic Stroke. Stroke 2018;49:e111–e122.

